# Why is non-suicidal self-injury more prevalent among women? Mediation and moderation analyses of psychological distress, emotion dysregulation, and impulsivity

**DOI:** 10.1101/2021.06.01.21258154

**Authors:** Nina M. Lutz, Sharon A.S. Neufeld, Roxanne W. Hook, Peter B. Jones, Ed T. Bullmore, Ian M. Goodyer, Samuel R. Chamberlain, Paul O. Wilkinson

## Abstract

**Objective:** Non-suicidal self-injury (NSSI) appears to be more common among women than men, though the underlying reasons for this remain unclear. In a community sample of young adults (*n*=996, aged 18-30) assessed during the COVID-19 pandemic, we investigated gendered patterns in NSSI etiology.

**Methods:** Mediation and moderation analyses considered associations between past-year NSSI prevalence, gender, and putative mechanistic variables: self-reported psychological distress (K10), emotion dysregulation (DERS), and impulsivity (UPPS-P).

**Results:** Nearly twice as many women as men reported past-year NSSI (14.47% versus 7.78%). Women reported significantly higher psychological distress and significantly lower sensation seeking and positive urgency than men. Psychological distress partially statistically mediated the relationship between gender and past-year NSSI. Gender did not significantly moderate associations between self-reported distress, emotion dysregulation, or impulsivity and past-year NSSI. Past-year NSSI prevalence did not significantly decrease with age and we found no significant age by gender interaction.

**Conclusions:** Greater levels of NSSI in young women are explained by their greater levels of emotional distress. Women do not appear to be more likely than men to report NSSI due to differences in how they manage emotional distress: gender did not moderate the association between psychological distress and past-year NSSI, and there were no gender differences in emotion dysregulation or negative urgency. Furthermore, we show that NSSI remains prevalent beyond adolescence. Early interventions which reduce distress or improve distress tolerance, strengthen emotion regulation skills, and provide alternative coping strategies merit investigation for NSSI.

**Highlights:**

- Young women were significantly more likely to report past-year NSSI than young men
- Psychological distress partially mediated the relationship between gender and NSSI
- Gender did not moderate associations between putative mechanistic variables and NSSI

## Introduction

Non-suicidal self-injury (NSSI) is the direct and deliberate damage of the body without suicidal intent (International Society for the Study of Self-Injury, 2018). Most often used to regulate emotions, NSSI may also be driven by a range of motivations such as self-punishment or communicating distress (Taylor et al., 2018). Broadly, NSSI can be considered a coping strategy which, though harmful, serves specific functions which individuals are able to rationally articulate (Lockwood et al., 2020). NSSI often begins between ages 12 and 14 (Cipriano et al., 2017) and prevalence appears to peak in mid-adolescence before declining into young adulthood (Moran et al., 2012; Plener et al., 2015), though both continuation and new initiation of NSSI remain common in young adults (Daukantaitė et al., 2020; Kiekens et al., 2019).

NSSI appears to be more common in women than men, with the largest gender differences emerging in clinical samples and among those with long-term repetitive NSSI (Bresin & Schoenleber, 2015; Daukantaitė et al., 2020). Past research reveals patterns of gender differences in method and location of self-injury (Sornberger et al., 2012), as well as motives for NSSI (Victor et al., 2018; Whitlock et al., 2011). Whilst the underlying reasons for these differences have received little attention in the literature, the study of psychological distress, emotion dysregulation, and impulsivity may prove fruitful.

Psychological distress, encompassing depressive and anxious symptomology (Kessler et al., 2002), has shown consistent longitudinal associations with NSSI in adolescents (Valencia-Agudo et al., 2018). When construed as a transdiagnostic latent factor across multiple symptoms of differing types of mental disorders, psychological distress also mediates the persistence of NSSI in adolescents and young adults (Polek et al., 2020). In contrast, emotion regulation encompasses the automatic and purposeful processes of managing emotional responses to external events (Gratz & Roemer, 2004). Poor emotion regulation skills have been robustly associated with NSSI risk (Wolff et al., 2019).

NSSI is often carried out with little planning (Nock & Prinstein, 2005), leading researchers to consider impulsivity an important risk factor. Impulsivity is multifaceted (Smith et al., 2007) and empirical support for the roles of disparate impulsive dimensions in NSSI is mixed, though it appears that self-report measures of mood-sensitive trait impulsivity are especially relevant (Hamza et al., 2015; Lockwood et al., 2017), with such measures prospectively predicting NSSI one year later (Cassels et al., 2020). Trouble sticking with difficult tasks and low planning have been hypothesized as specifically associated with recurrent NSSI even in those who want to stop (Lockwood et al., 2017; Riley et al., 2015); cessation as a long-term goal requires persevering through unpleasant feelings and urges to perform NSSI whilst committing to alternative coping strategies.

Recent qualitative work by Lockwood and colleagues (2020) describes how psychological distress, emotion dysregulation, and impulsivity each contribute to self-harm (either explicitly non-suicidal or unclear/mixed suicidal motive). Most participants describe feeling unable to manage distress which escalates until reaching a “tipping point” leading to NSSI. Many are very aware of the negative consequences of their self-harm, but these cannot outweigh the strength of the urge or the emotional relief brought by the act. Most also describe acting on impulse at least some of the time, hurting themselves “out of the blue” without conscious awareness of why they are self-harming (Lockwood et al., 2020). Though other distal and proximal risk factors are undoubtedly involved (Fox et al., 2015), these three variables – psychological distress, emotion dysregulation, and impulsivity – appear to be particularly salient in understanding NSSI.

In the present community sample of young adults, we hypothesized that past-year NSSI would be more prevalent among women than men. If this was shown to be the case, we then planned to test two sets of competing (though not mutually exclusive) hypotheses to explain the gender gap. First, psychological distress, emotion dysregulation, or impulsivity may *mediate* the relationship between gender and past-year NSSI; the NSSI gender gap may be accounted for by different levels of these traits in women versus men. Second, gender may *moderate* the effects of psychological distress, emotion dysregulation, or impulsivity on past-year NSSI; these traits may be more strongly related to NSSI in women than in men. Mediation and moderation analyses both help us understand the relationship between an independent and dependent variable: mediators explain how the two variables are related, while moderators affect the strength and direction of that association.

In addition to these primary analyses, we hypothesized that prevalence of past-year NSSI would decrease over ages 18 to 33 (Daukantaitė et al., 2020; Moran et al., 2012; Plener et al., 2015), and we planned to examine the interaction between age and gender to ascertain whether age moderates the gender difference in past-year NSSI prevalence across young adulthood.

## Methods

### Participants

Participants were *n*=996 young adults aged 18 to 33 years (*M*=25.54, *SD*=3.10), 63.15% (*n*=629) identifying as female. Data were collected May-July 2020 as part of a study assessing mental health during the COVID-19 pandemic within the NeuroScience in Psychiatry Network (NSPN) cohort. The NSPN cohort is a general population sample of *n*=2403 individuals across Cambridgeshire and Greater London, aged 14 to 24 at baseline, established in 2012 to longitudinally investigate both healthy and psychopathological development. NSPN participants were originally recruited via invitations sent by primary care physicians, schools and Further Education colleges, and by purposive advertisement. Baseline cohort characteristics, as well as previous follow-up data, have been previously described elsewhere (Kiddle et al., 2018).

To recruit the present sample, emails were sent to all *n*=2036 participants who had not withdrawn from NSPN. Responses were received from *n*=1005. An additional *n*=22 participants from a separate NSPN depression cohort were invited, for an initial sample of *n*=1027. Of the *n*=1027, *n*=6 returned submissions with no responses; *n*=5 reported an age inconsistent with baseline NSPN data, suggesting the survey had been completed by a parent (i.e., participant may have provided their parents’ contact details rather than their own); a further *n*=19 did not respond to the NSSI measure, and *n*=1 did not report age or gender. Thus, we had a final sample of *n*=996, of whom *n*=21 came from the depression cohort.

Compared to NSPN participants who did not participate in this follow-up, those who did were significantly older (*p*=0.004) and more likely to identify as female (*p*<0.001). There were no differences in ethnicity (*p*=0.53), baseline levels of psychological distress (*p*=0.56), or baseline prevalence of past-year NSSI (*p*=0.09). The majority of participants in the present sample (71.29%) have attained an undergraduate degree, postgraduate degree, or professional qualification. In the present sample, 78.01% reported their ethnicity as White, 9.84% Asian, 3.82% Black, 6.33% mixed or multiple ethnic groups, and 1.41% another ethnicity. These rates are comparable to the baseline NSPN cohort, which is broadly representative of ethnicity in the general population of England and Wales (Kiddle et al., 2018).

Ethical approval for the current study was provided by the Cambridge East Research Ethics Committee (reference number 16/EE/0260, project: *Intermediate phenotypes of impulsivity and compulsivity*, Chief Investigator SRC) following the Declaration of Helsinki. Participants provided electronic informed consent and were compensated £25.

### Materials

#### Drugs, Alcohol and Self-Injury Questionnaire (DASI)

NSSI history was assessed via a yes/no question from the DASI, with established validity and reliability (Wilkinson et al., 2018): “*Have you ever tried to hurt yourself on purpose without trying to kill yourself? (for example: things like burning, cutting or scratching yourself)*.*”* Participants who endorsed lifetime NSSI were then asked how many times they had hurt themselves in the past month, past year, since turning 18, and in their entire life. Frequency response options for each timeframe were: never, once, 2-4 times, 5-10 times, more than 10 times.

#### Kessler Psychological Distress Scale (K10)

The K10 includes ten questions assessing past-month anxious and depressive symptoms (e.g. feeling nervous, hopeless, tired, restless, sad, worthless) (Kessler et al., 2002). It demonstrates consistent psychometric properties across women and men and can therefore be used to compare trait gender differences (Drapeau et al., 2010; Kessler et al., 2002).

#### Difficulties in Emotion Regulation Scale (DERS)

The 18-item short form generates six sub-scores capturing different components of trait emotion dysregulation. Here we utilize the DERS total score since all subscales are consistently associated with NSSI (Wolff et al., 2019). The DERS demonstrates strong psychometric properties (Kaufman et al., 2016) and measurement invariance across men and women (Ritschel et al., 2015).

#### UPPS-P

The 20-item short form contains five trait impulsive subscales (Cyders et al., 2014): *Negative Urgency*, the tendency to react rashly to negative emotions; *Lack of Perseverance*, a difficulty seeing thing through to the end; *Lack of Premeditation*, the tendency to act without thinking; *Sensation Seeking*, the tendency to pursue novel and thrilling experiences; and *Positive Urgency*, the tendency to react rashly to positive emotions. The UPPS-P demonstrates good construct validity (Smith et al., 2007) and measurement invariance across men and women (Cyders, 2013).

### Statistical Analysis

All analyses were conducted in STATA 14.2. Age and gender distributions between those with and without past-year NSSI were compared using a t-test and Fisher’s Exact Test. Seven participants who reported their gender identity as “other” were removed from primary analyses investigating whether putative mechanistic variables [psychological distress (K10 score), emotion dysregulation (DERS total score), or impulsivity (UPPS-P subscales)] contribute to the past-year NSSI gender gap. Univariate binary logistic regressions related each self-report score to NSSI (yes versus no past-year NSSI) and gender (female versus male identifying). Variables associated with both NSSI and gender were investigated as possible statistical mediators between gender and NSSI, using bootstrapped generalized structural equation models. Variables significantly associated with NSSI were included in binary logistic regression moderation analyses, with an interaction term between gender and each self-report measure.

Two multiple binary logistic regressions analyzed linear and quadratic interaction effects of age and gender on past-year NSSI. For descriptive purposes, age bins are used in Figure 2; however age was a continuous variable in regression analyses.

**Figure 1.**
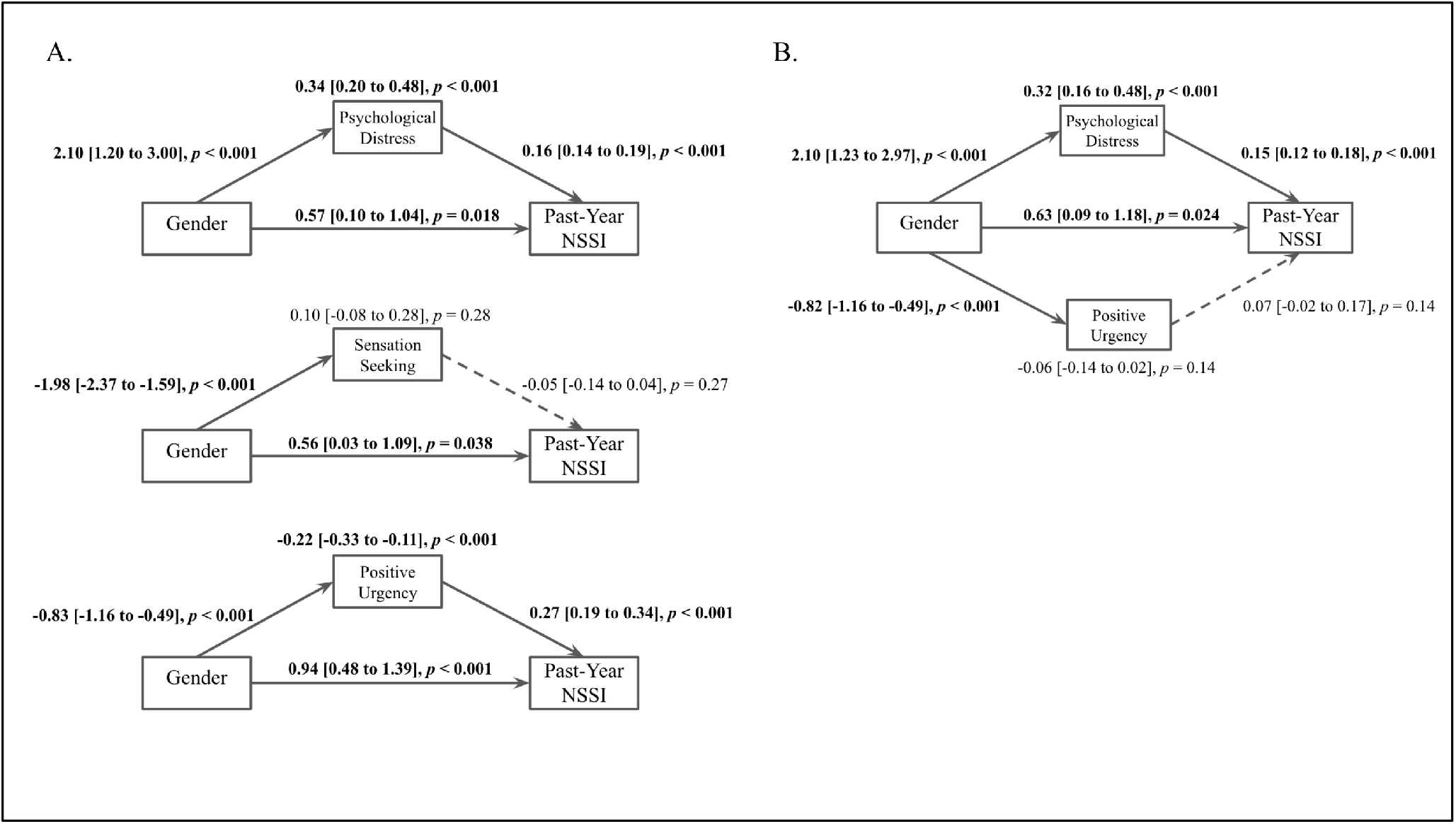
Path diagrams of the effect of gender (female or male) on past-year NSSI via psychological distress, sensation seeking, and positive urgency individually (A) and in a joint model including psychological distress and positive urgency (B). The models show standardized coefficients with 95% confidence intervals of direct effects between variables (displayed adjacent to each respective arrow) and indirect effects of gender on NSSI through each mediator (displayed above respective mediators). Bold text and solid lines indicate significance at *p*<0.05.

**Figure 2.**
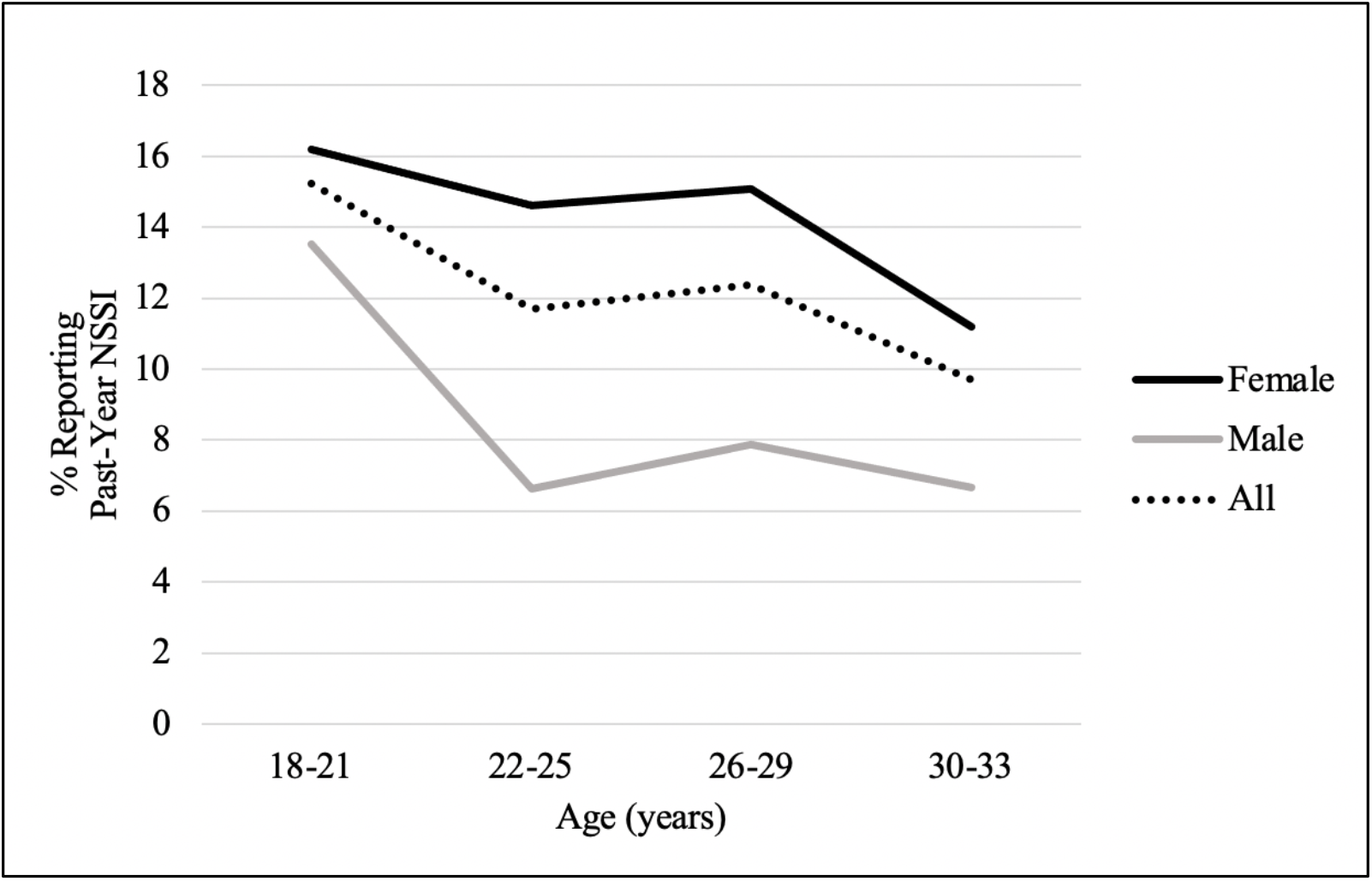
Effects of age and gender on prevalence of past-year NSSI

## Results

### Descriptive Statistics

Overall, 31.12% of the sample (*n*=331) reported lifetime NSSI and 12.55% (*n*=125) reported past-year NSSI. Participants with or without past-year NSSI did not differ in age (t(994)=1.17, *p*=0.24). Gender distribution significantly differed by group (Fisher’s Exact Test, *p*<0.001): 14.47% of women and 7.78% of men reported past-year NSSI, as did six of the seven participants (85.71%) who reported their gender identity as “other”.

### Univariate Associations of Putative Mechanistic Variables with NSSI and Gender

Univariate regressions (Table 1) showed the following significant associations with past-year NSSI: greater psychological distress, emotion dysregulation, negative urgency, lack of premeditation, and positive urgency, as well as lower sensation seeking. Female gender was significantly associated with greater psychological distress, while male gender was associated with greater sensation seeking and positive urgency. These three variables, associated with both gender and NSSI, were considered in the following mediation analyses.

**Table 1.** Univariate associations between gender/past-year NSSI and putative mechanistic variables. Bold indicates significance at *p*<0.05.

|  | Gender |  |  |  | Past-Year NSSI |  |  |  |
| --- | --- | --- | --- | --- | --- | --- | --- | --- |
|  | Women<br>*M(SD)* | Men<br>*M(SD)* | Coef.<br>[95% CI] | *p* | Yes<br>*M(SD)* | No<br>*M(SD)* | Coef.<br>[95% CI] | *p* |
| Psychological Distress<sup>a</sup> | 22.54<br>(7.40) | 20.44<br>(7.59) | 2.10<br>[1.13, 3.07] | **<0.001** | 29.94<br>(7.64) | 20.65<br>(6.80) | 0.16<br>[0.13, 0.19] | **<0.001** |
| Emotion Dysregulation<sup>b</sup> | 38.33<br>(11.67) | 36.82<br>(11.82) | 1.51<br>[-0.09, 3.11] | 0.065 | 49.59<br>(12.62) | 36.02<br>(10.56) | 0.09<br>[0.07, 0.11] | **<0.001** |
| Negative Urgency<sup>c</sup> | 8.60<br>(2.68) | 8.68<br>(2.56) | -0.08<br>[-0.43, 0.26] | 0.64 | 10.68<br>(2.54) | 8.35<br>(2.52) | 0.36<br>[0.27, 0.44] | **<0.001** |
| Lack of Perseverance<sup>c</sup> | 8.38<br>(1.80) | 8.25<br>(1.71) | 0.13<br>[-0.10, 0.36] | 0.28 | 8.30<br>(2.00) | 8.34<br>(1.74) | -0.01<br>[-0.12, 0.10] | 0.81 |
| Lack of Premeditation<sup>c</sup> | 7.89<br>(0.07) | 7.85<br>(0.10) | 0.04<br>[-0.20, 0.27] | 0.75 | 8.41<br>(1.83) | 7.81<br>(1.77) | 0.19<br>[0.08, 0.29] | **0.001** |
| Sensation Seeking<sup>c</sup> | 8.91<br>(2.50) | 10.89<br>(2.50) | -1.98<br>[-2.30, -1.65] | **<0.001** | 9.12<br>(2.17) | 9.70<br>(2.66) | -0.08<br>[-0.16, -0.01] | **0.029** |
| Positive Urgency<sup>c</sup> | 6.77<br>(2.17) | 7.59<br>(2.41) | -0.83<br>[-1.12, -0.53] | **<0.001** | 8.22<br>(2.59) | 6.91<br>(2.21) | 0.23<br>[0.15, 0.31] | **<0.001** |

<sup>a</sup>Kessler Psychological Distress Scale (K10)
<sup>b</sup>Difficulties in Emotion Regulation Scale (DERS) short form
<sup>c</sup>UPPS-P impulsivity questionnaire short form

### Mediation Analyses

Psychological distress significantly mediated the relationship between gender and past-year NSSI (Figure 1): 37% of the effect of gender on past-year NSSI was explained via psychological distress. Sensation seeking was not a significant mediator. Positive urgency negatively mediated the effect of gender on past-year NSSI, since men reported greater positive urgency than women, and this urgency was related to increased NSSI. In a joint mediation model, only psychological distress showed a significant effect.

### Moderation Analyses

All analyses were non-significant: gender did not significantly moderate the effects of psychological distress (*b*=0.002, 95%CI[-0.06,0.07], *p*=0.95), emotion dysregulation (*b*=-0.007, 95%CI[-0.05,0.03], *p*=0.74), negative urgency (*b*=-0.03, 95%CI[-0.21,0.16], *p*=0.77), lack of premeditation (*b*=-0.08, 95%CI[-0.15,0.32], *p*=0.50), sensation seeking (*b*=0.05, 95%CI[-0.13,0.22], *p*=0.61), or positive urgency (*b*=0.03, 95%CI[-0.15,0.21], *p*=0.76) on past-year NSSI.

### Age Effects, and Their Moderation by Gender

Past-year NSSI prevalence was lower at older ages, but this was not statistically significant. The gender difference did not change with age (linear and quadratic age x gender interactions non-significant; Table 2, Figure 2).

**Table 2.** Age and gender effects on past-year NSSI prevalence

|  | OR | 95% CI | *p* |
| --- | --- | --- | --- |
| Gender | 2.00 | 1.29, 3.13 | **0.002** |
| Age | 0.96 | 0.90, 1.02 | 0.20 |
| Age<sup>2</sup> | 1.00 | 0.98, 1.02 | 0.91 |
| Age x Gender | 1.02 | 0.88, 1.18 | 0.80 |
| Age<sup>2</sup> x Gender | 1.00 | 0.96, 1.05 | 0.98 |

### Comparison to NSSI prevalence pre-COVID-19

Given the timing of data collection, high NSSI rates could be related to worsening mental health during the COVID-19 pandemic (Veldhuis et al., 2021), and so may not represent the population at normal times. We tested for this in a secondary analysis: the majority of participants (*n*=548) reported on NSSI in a previous follow-up study of the NSPN cohort in 2017-2018 (Chamberlain et al., 2019). Those data were collected when participants were aged 17-31 (*M*=23.42 years), an average of 28.81 months prior to the data presented here. Past-year NSSI prevalence did not differ significantly between the two time points (McNemar’s Test, χ^2^(1, *n*=548)=2.85, *p*=0.09; 13.50% for pre-pandemic versus 10.77% for intra-pandemic).

## Discussion

In this community sample of young adults, we investigated how psychological distress, emotion dysregulation, and impulsivity may contribute to a higher prevalence of NSSI among women. Our primary finding was that psychological distress statistically mediated the relationship between gender and past-year NSSI - women were twice as likely as men to report past-year NSSI, with over a third of the variance of NSSI in women explained by their significantly higher levels of distress. This then leads to the question: *why* are young women reporting worse mental health than young men?

Studies have consistently shown that women display significantly higher psychological distress than men (Drapeau et al., 2010; Matud et al., 2015; Nurullah, 2010; Seedat et al., 2009). Some researchers suggest this is because questionnaire items are biased towards “feminine” expressions of distress, and women are socialized to more freely self-disclose mental health symptoms (Nurullah, 2010). However, psychometric studies establish that men and women respond to items similarly on the K10 (Drapeau et al., 2010; Kessler et al., 2002), so it is unlikely that the present effect is due to gender bias in the assessment tool. Rather, higher distress among women may be linked to greater exposure to life stressors, as well as gender differences in coping styles and psychological resources (Nurullah, 2010). As our analysis is cross-sectional, we cannot establish the direction of causality and so it is possible that higher levels of NSSI may itself lead to greater psychological distress among women. Indeed, NSSI is bi-directionally related to distress (Buelens et al., 2019) including in this cohort studied prior to the COVID-19 pandemic (Polek et al., 2020). NSSI can elicit feelings of shame, guilt, and self-criticism (Bachtelle & Pepper, 2015; Daly & Willoughby, 2019) and strain interpersonal relationships (Waals et al., 2018), with many people who engage in NSSI reporting feeling unsupported by family, friends, and mental health professionals (Tillman et al., 2018).

The findings also note a significant negative statistically mediating effect of positive urgency on NSSI. Positive urgency likely weakened the overall association between gender and NSSI because men displayed higher positive urgency, which was associated with NSSI – yet women were more likely to report NSSI. Recent research has begun to investigate the relationship between gender, positive urgency, and NSSI (Peckham et al., 2020) based on evidence that men report higher positive urgency than women (Cyders, 2013) and more risky behaviors while experiencing positive affect (Cyders & Smith, 2010). Men are also more likely to report NSSI which seems conceptually related to positive urgency, e.g. hurting themselves as part of a game with friends (Laye-Gindhu & Schonert-Reichl, 2005). As Bresin and Schoenleber (2015) point out, “NSSI might be part of a broader class of dangerous, thrill-seeking, but socially acceptable, behaviors that are common for men” (p. 60).

We found no evidence that gender moderated the associations between NSSI and psychological distress, emotion dysregulation, negative urgency, lack of premeditation, positive urgency, or sensation seeking – these measures were similarly associated with past-year NSSI in women and men. This finding is consistent with research showing that UPPS-P subscales are similarly associated with risky behaviors across genders (Cyders, 2013), and gender does not moderate the relationships between emotion dysregulation, positive urgency, or negative urgency and NSSI (Peckham et al., 2020; Wolff et al., 2019).

Also in line with past research, we found that NSSI was cross-sectionally associated with higher psychological distress (Valencia-Agudo et al., 2018), emotion dysregulation (Wolff et al., 2019), positive urgency (Claes et al., 2015; Claes & Muehlenkamp, 2013), negative urgency, and lack of premeditation (Hamza et al., 2015). However, in contrast to some studies (Hamza et al., 2015; Lockwood et al., 2017; Riley et al., 2015), NSSI was not associated with lack of perseverance and was significantly associated with lower, rather than higher, sensation seeking. Additional research relating these traits to aspects of NSSI (e.g., NSSI purpose or methods) could clarify whether they are primarily relevant to certain presentations of NSSI.

Age did not moderate the gender difference in past-year NSSI prevalence. Prevalence did not significantly decrease with age, and women and men did not differ in their pattern of past-year NSSI endorsement across young adulthood. Longitudinal studies and meta-analyses show that endorsement of NSSI typically declines beyond teenage years (Daukantaitė et al., 2020; Moran et al., 2012; Swannell et al., 2014), leading to assertions that “most self-harming behavior in adolescents resolves spontaneously” (Moran et al., 2012). However, both lifetime and past-year NSSI rates in the present sample are substantially higher than those reported in past studies of young adults (Moran et al., 2012; Swannell et al., 2014; Taliaferro & Muehlenkamp, 2015) - the true scope of NSSI among young adults, and the reason for discrepancies, remains unclear.

It is possible that some participants sampled here had previously ceased NSSI but returned to this behavior to cope with worsening mental health during the COVID-19 pandemic. Though the magnitude of the pandemic’s impact on mental health is difficult to determine, initial evidence shows elevated psychological distress and worsening psychiatric symptoms across both clinical and community samples (Veldhuis et al., 2021; Vindegaard & Benros, 2020; Wang et al., 2020), and experts have expressed concern that the reduction of face-to-face mental health services and accumulation of risk factors will lead to a “steep increase in NSSI” (Plener, 2021). However, we found no evidence of such an increase in this cohort – compared to data collected during a separate NSPN cohort follow-up study in 2017-2018, past-year NSSI prevalence here was numerically (though not statistically significantly) lower. Longitudinal research examining the impact of the pandemic on NSSI is needed.

### Limitations

The data reported here are cross-sectional and so we cannot demonstrate true mediation, but rather statistical mediation. Longitudinal associations between NSSI and the variables considered here are likely complex and bi-directional, as shown in analyses of previous waves of NSPN data reported elsewhere (Cassels et al., 2020; Polek et al., 2020). Thus, it is possible that NSSI mediated the effects of gender on distress, rather than the other way around. The non-significant association between age and prevalence, despite rates appearing to decline (Figure 2), may mean that analyses were underpowered to detect a significant effect. Also, our NSSI measure did not delineate engagement before versus after the start of the COVID-19 pandemic.

This study is also limited by our binary classification of gender. We do not know how many participants are transgender, and analyses excluded the seven participants who reported their gender identity as “other”, all of whom reported lifetime NSSI. This is, unfortunately, consistent with much of the NSSI literature; studies of gender differences have binarized gender as male/female, either failing to ask participants whether they are transgender or excluding non-cisgender participants from analyses (Bresin & Schoenleber, 2015; Victor et al., 2018). This is despite mounting evidence that NSSI is substantially more prevalent among trans and gender non-conforming people (Liu et al., 2019; Marshall et al., 2016).

Gendered assumptions about NSSI likely result in chronic underreporting among men (Green & Jakupcak, 2015; Kimbrel et al., 2017). Men often self-injure by punching walls or objects (Whitlock et al., 2011), yet the majority of research has not included this as an NSSI method (Kimbrel et al., 2017). This bias in NSSI measurement can have substantial impact; in a predominantly male sample, endorsement of lifetime NSSI increased by 20% when wall/object punching was explicitly included (Kimbrel et al., 2018). The measure used here only mentions burning, cutting, and scratching as examples of NSSI. It is therefore possible that we have underreported the true prevalence of NSSI among men in this sample.

### Future directions

Compared to studies of adolescents, relatively few have focused on NSSI in young adulthood. The relatively high rates of NSSI reported in the present study highlight the need for additional research. Given developmental changes from adolescence to young adulthood, researchers cannot assume the adolescent literature will generalize to older populations. There is particular need for research on differences between adolescent and adult onset NSSI, and longitudinal studies examining the course of this behavior. Research should be sensitive to the ways that NSSI may be experienced by people of different genders, including gender non-conforming identities. The intention should not be to define “male NSSI” and “female NSSI”. Rather, by considering gendered patterns in the etiology and presentation of NSSI, we may improve our understanding of the complex interplay of individual, social, environmental, and biological factors which shape mental health.

Future research should consider variables which affect the relationships between risk factors and NSSI. The literature on mediators and moderators is still limited and almost exclusively focused on adolescents, with many studies either revealing conflicting results or needing replication (Valencia-Agudo et al., 2018). Furthermore, beyond identifying correlates of NSSI, the literature should investigate how etiologic factors relate to the experience of NSSI. A recent study by Lockwood and colleagues (2020) is one such example, utilizing qualitative methods to explore the cognitive and emotional factors involved in the build-up to acts of self-harm. The researchers demonstrate that people who engage in self-harm can offer valuable insights into the processes underlying their behavior and underscore that there is wide variation in NSSI experience both within and between individuals.

Clinicians should be aware of how common NSSI is among young adults. Findings here reveal potential avenues for early interventions, including developing strategies which reduce psychological distress or improve distress tolerance, strengthen emotion regulation skills and control impulsive reactions, and replace NSSI with healthier coping mechanisms. This parallels meta-analytic findings that Dialectical Behavioral Therapy, which emphasizes these skills, effectively reduces self-harm in adolescents (Kothgassner et al., 2020).

### Conclusion

Our results demonstrate that young women experience more psychological distress than young men, and this difference in mental health significantly contributes to the substantial gender gap in past-year NSSI prevalence. However, we found no evidence that the effects of distress on NSSI are different between women and men. NSSI often persists into young adulthood and should not be viewed as a teenage behavior, likely to resolve on its own. The high prevalence of self-injury in this sample speaks to the urgent need for clinical interventions which effectively reduce NSSI.

## Data Availability

Data are available upon reasonable request to the corresponding author

## Acknowledgements

We sincerely thank the NeuroScience in Psychiatry Network (NSPN) study team and the study participants for their contribution to this research.

## References

Bachtelle, S. E., & Pepper, C. M. (2015). The Physical Results of Nonsuicidal Self-Injury: The Meaning behind the Scars. Journal of Nervous and Mental Disease, 203(12), 927–933. https://doi.org/10.1097/NMD.0000000000000398

Bresin, K., & Schoenleber, M. (2015). Gender differences in the prevalence of nonsuicidal self-injury: A meta-analysis. Clinical Psychology Review, 38, 55--64. https://doi.org/10.1016/j.cpr.2015.02.009

Buelens, T., Luyckx, K., Gandhi, A., Kiekens, G., & Claes, L. (2019). Non-Suicidal Self-Injury in Adolescence: Longitudinal Associations with Psychological Distress and Rumination. Journal of Abnormal Child Psychology, 47(9), 1569–1581. https://doi.org/10.1007/s10802-019-00531-8

Cassels, M., Neufeld, S., van Harmelen, A. L., Goodyer, I., & Wilkinson, P. (2020). Prospective pathways from impulsivity to non-suicidal self-injury among youth. Archives of Suicide Research.

Chamberlain, S. R., Tiego, J., Fontenelle, L. F., Hook, R., Parkes, L., Segrave, R., Hauser, T. U., Dolan, R. J., Goodyer, I. M., Bullmore, E., Grant, J. E., & Yücel, M. (2019). Fractionation of impulsive and compulsive trans-diagnostic phenotypes and their longitudinal associations. Australian and New Zealand Journal of Psychiatry, 53(9), 896–907. https://doi.org/10.1177/0004867419844325

Cipriano, A., Cella, S., & Cotrufo, P. (2017). Nonsuicidal self-injury: A systematic review. Frontiers in Psychology, 8, 1946. https://doi.org/10.3389/fpsyg.2017.01946

Claes, L., Islam, M. A., Fagundo, A. B., Jimenez-Murcia, S., Granero, R., Agüera, Z., Rossi, E., Menchón, J. M., & Fernández-Aranda, F. (2015). The relationship between non-suicidal self-injury and the UPPS-P impulsivity facets in eating disorders and healthy controls. PLoS ONE, 10(5), 1–11. https://doi.org/10.1371/journal.pone.0126083

Claes, L., & Muehlenkamp, J. (2013). The Relationship between the UPPS-P Impulsivity Dimensions and Nonsuicidal Self-Injury Characteristics in Male and Female High-School Students. Psychiatry Journal, 2013, 654847. https://doi.org/10.1155/2013/654847

Cyders, M. A. (2013). Impulsivity and the Sexes: Measurement and Structural Invariance of the UPPS-P Impulsive Behavior Scale. Assessment, 20(1), 86–97. https://doi.org/10.1177/1073191111428762

Cyders, M. A., Littlefield, A. K., Coffey, S., & Karyadi, K. A. (2014). Examination of a short English version of the UPPS-P Impulsive Behavior Scale. Addictive Behaviors, 39(9), 1372–1376. https://doi.org/10.1016/j.addbeh.2014.02.013

Cyders, M. A., & Smith, G. T. (2010). Longitudinal validation of the urgency traits over the first year of college. Journal of Personality Assessment, 92(1), 63–69. https://doi.org/10.1080/00223890903381825

Daly, O., & Willoughby, T. (2019). A longitudinal study investigating bidirectionality among nonsuicidal self-injury, self-criticism, and parental criticism. Psychiatry Research, 271, 678–683. https://doi.org/10.1016/j.psychres.2018.12.056

Daukantaite, D., Lundh, L.-G., Wångby-Lundh, M., Claréus, B., Bjärehed, J., Zhou, Y., & Liljedahl, S. I. (2020). What happens to young adults who have engaged in self-injurious behavior as adolescents? A 10-year follow-up. European Child & Adolescent Psychiatry. https://doi.org/10.1007/s00787-020-01533-4

Drapeau, A., Beaulieu-Prévost, D., Marchand, A., Boyer, R., Préville, M., & Kairouz, S. (2010). A life-course and time perspective on the construct validity of psychological distress in women and men. Measurement invariance of the K6 across gender. BMC Medical Research Methodology, 10, 68. https://doi.org/10.1186/1471-2288-10-68

Fox, K. R., Franklin, J. C., Ribeiro, J. D., Kleiman, E. M., Bentley, K. H., & Nock, M. K. (2015). Meta-analysis of risk factors for nonsuicidal self-injury. Clinical Psychology Review, 42, 156–167. https://doi.org/10.1016/j.cpr.2015.09.002

Gratz, K. L., & Roemer, L. (2004). Multidimensional assessment of emotion regulation and dysregulation. Journal of Psychopathology and Behavioral Assessment, 26(1), 41–54. https://doi.org/10.1023/B:JOBA.0000007455.08539.94

Green, J. D., & Jakupcak, M. (2015). Masculinity and men’s self-harm behaviors: Implications for Non-Suicidal Self-Injury Disorder. Psychology of Men and Masculinity, 17(2), 147–155. https://doi.org/10.1037/a0039691

Hamza, C. A., Willoughby, T., & Heffer, T. (2015). Impulsivity and nonsuicidal self-injury: A review and meta-analysis. Clinical Psychology Review, 38, 13–24. https://doi.org/10.1016/j.cpr.2015.02.010

International Society for the Study of Self-Injury. (2018). What is self-injury? https://itriples.org/about-self-injury/what-is-self-injury

Kaufman, E. A., Xia, M., Fosco, G., Yaptangco, M., Skidmore, C. R., & Crowell, S. E. (2016). The Difficulties in Emotion Regulation Scale Short Form (DERS-SF): Validation and Replication in Adolescent and Adult Samples. Journal of Psychopathology and Behavioral Assessment, 38, 443–455. https://doi.org/10.1007/s10862-015-9529-3

Kessler, R. C., Andrews, G., Colpe, L. J., Hiripi, E., Mroczek, D. K., Normand, S. L. T., Walters, E. E., & Zaslavsky, A. M. (2002). Short screening scales to monitor population prevalences and trends in non-specific psychological distress. Psychological Medicine, 32, 959–976. https://doi.org/10.1017/S0033291702006074

Kiddle, B., Inkster, B., Prabhu, G., Moutoussis, M., Whitaker, K. J., Bullmore, E. T., Dolan, R. J., Fonagy, P., Goodyer, I. M., Jones, P. B., Dolan, R., Hauser, T., Vértes, P., Whitaker, K., Villis, L., Bhatti, J., Ooi, C., Widmer, B., Alrumaithi, A., … van Harmelen, A. L. (2018). Cohort profile: The NSPN 2400 Cohort: A developmental sample supporting the Wellcome Trust Neuro Science in Psychiatry Network. International Journal of Epidemiology, 47(1), 18–19g. https://doi.org/10.1093/ije/dyx117

Kiekens, G., Hasking, P., Claes, L., Boyes, M., Mortier, P., Auerbach, R. P., Cuijpers, P., Demyttenaere, K., Green, J. G., Kessler, R. C., Myin-Germeys, I., Nock, M. K., & Bruffaerts, R. (2019). Predicting the incidence of non-suicidal self-injury in college students. European Psychiatry, 59, 44–51. https://doi.org/10.1016/j.eurpsy.2019.04.002

Kimbrel, N. A., Calhoun, P. S., & Beckham, J. C. (2017). Nonsuicidal self-injury in men: a serious problem that has been overlooked for too long. World Psychiatry, 16(1), 108–109. https://doi.org/10.1002/wps.20358

Kimbrel, N. A., Thomas, S. P., Hicks, T. A., Hertzberg, M. A., Clancy, C. P., Elbogen, E. B., Meyer, E. C., DeBeer, B. B., Gross, G. M., Silvia, P. J., Morissette, S. B., Gratz, K. L., Calhoun, P. S., & Beckham, J. C. (2018). Wall/Object Punching: An Important but Under-Recognized Form of Nonsuicidal Self-Injury. Suicide and Life-Threatening Behavior, 48(5), 501–511. https://doi.org/10.1111/sltb.12371

Kothgassner, O., D., Robinson, K., Goreis, A., Ougrin D, & Plener, P. L. (2020). Does treatment method matter? A meta-analysis of the past 20 years of research on therapeutic interventions for self-harm and suicidal ideation in adolescents. Borderline Personality Disorder and Emotion Dysregulation, 7, 9. https://doi.org/10.1186/s40479-020-00123-9

Laye-Gindhu, A., & Schonert-Reichl, K. A. (2005). Nonsuicidal self-harm among community adolescents: Understanding the “whats” and “whys” of self-harm. Journal of Youth and Adolescence, 34(5), 447–457. https://doi.org/10.1007/s10964-005-7262-z

Liu, R. T., Sheehan, A. E., Walsh, R. F. L., Sanzari, C. M., Cheek, S. M., & Hernandez, E. M. (2019). Prevalence and correlates of non-suicidal self-injury among lesbian, gay, bisexual, and transgender individuals: A systematic review and meta-analysis. Clinical Psychology Review, 74, 101783. https://doi.org/10.1016/j.cpr.2019.101783

Lockwood, J., Daley, D., Townsend, E., & Sayal, K. (2017). Impulsivity and self-harm in adolescence: a systematic review. European Child and Adolescent Psychiatry, 26(4), 387– 402. https://doi.org/10.1007/s00787-016-0915-5

Lockwood, J., Townsend, E., Allen, H., Daley, D., & Sayal, K. (2020). What young people say about impulsivity in the short-term build up to self-harm: A qualitative study using card-sort tasks. PLoS ONE, 15(12), e0244319.

Marshall, E., Claes, L., Bouman, W. P., Witcomb, G. L., & Arcelus, J. (2016). Non-suicidal self-injury and suicidality in trans people: A systematic review of the literature. International Review of Psychiatry, 28(1), 58–69. https://doi.org/10.3109/09540261.2015.1073143

Matud, M. P., Bethencourt, J. M., & Ibáñez, I. (2015). Gender differences in psychological distress in Spain. International Journal of Social Psychiatry, 61(6), 560–568. https://doi.org/10.1177/0020764014564801

Moran, P., Coffey, C., Romaniuk, H., Olsson, C., Borschmann, R., Carlin, J. B., & Patton, G. C. (2012). The natural history of self-harm from adolescence to young adulthood: A population-based cohort study. The Lancet, 379(9812), 236–243. https://doi.org/10.1016/S0140-6736(11)61141-0

Nock, M. K., & Prinstein, M. J. (2005). Contextual features and behavioral functions of self-mutilation among adolescents. Journal of Abnormal Psychology, 114(1), 140–146. https://doi.org/10.1037/0021-843X.114.1.140

Nurullah, A. S. (2010). Gender Differences in Distress: The Mediating Influence of Life Stressors and Psychological Resources. Asian Social Science, 6(5), 27–35. https://doi.org/10.5539/ass.v6n5p27

Peckham, A. D., Jordan, H., Silverman, A., Jarvi Steele, S., Björgvinsson, T., & Beard, C. (2020). From Urges to Action: Negative Urgency and Nonsuicidal Self-Injury in an Acute Transdiagnostic Sample. Archives of Suicide Research, 24, 367–383. https://doi.org/10.1080/13811118.2019.1625831

Plener, P. L. (2021). COVID-19 and Nonsuicidal Self-Injury: The Pandemic’s Influence on an Adolescent Epidemic. American Journal of Public Health, 111(2), 195–196. https://doi.org/10.2105/AJPH.2020.306037

Plener, P. L., Schumacher, T. S., Munz, L. M., & Groschwitz, R. C. (2015). The longitudinal course of non-suicidal self-injury and deliberate self-harm: a systematic review of the literature. Borderline Personality Disorder and Emotion Dysregulation, 2(1), 2. https://doi.org/10.1186/s40479-014-0024-3

Polek, E., Neufeld, S. A. S., Wilkinson, P., Goodyer, I., St Clair, M., Prabhu, G., Dolan, R., Bullmore, E. T., Fonagy, P., Stochl, J., & Jones, P. B. (2020). How do the prevalence and relative risk of non-suicidal self-injury and suicidal thoughts vary across the population distribution of common mental distress (the p factor)? Observational analyses replicated in two independent UK cohorts of young people. BMJ Open, 10, e032494. https://doi.org/10.1136/bmjopen-2019-032494

Riley, E. N., Combs, J. L., Jordan, C. E., & Smith, G. T. (2015). Negative Urgency and Lack of Perseverance: Identification of Differential Pathways of Onset and Maintenance Risk in the Longitudinal Prediction of Nonsuicidal Self-Injury. Behavior Therapy, 46(4), 439–448. https://doi.org/10.1016/j.beth.2015.03.002

Ritschel, L. A., Tone, E. B., Schoemann, A. M., & Lim, N. E. (2015). Psychometric properties of the difficulties in emotion regulation scale across demographic groups. Psychological Assessment, 27(3), 944–954. https://doi.org/10.1037/pas0000099

Seedat, S., Scott, K. M., Angermeyer, M. C., Berglund, P., Bromet, E. J., Brugha, T. S., Demyttenaere, K., De Girolamo, G., Haro, J. M., Jin, R., Karam, E. G., Kovess-Masfety, V., Levinson, D., Medina Mora, M. E., Ono, Y., Ormel, J., Pennell, B. E., Posada-Villa, J., Sampson, N. A., … Kessler, R. C. (2009). Cross-national associations between gender and mental disorders in the World Health Organization World Mental Health Surveys. Archives of General Psychiatry, 66(7), 785–795. https://doi.org/10.1001/archgenpsychiatry.2009.36

Smith, G. T., Fischer, S., Cyders, M. A., Annus, A. M., Spillane, N. S., & McCarthy, D. M. (2007). On the validity and utility of discriminating among impulsivity-like traits. Assessment, 14(2), 155–170. https://doi.org/10.1177/1073191106295527

Sornberger, M. J., Heath, N. L., Toste, J. R., & McLouth, R. (2012). Nonsuicidal self-injury and gender: Patterns of prevalence, methods, and locations among adolescents. Suicide and Life-Threatening Behavior, 42(3), 266–278. https://doi.org/10.1111/j.1943-278X.2012.0088.x

Swannell, S. V., Martin, G. E., Page, A., Hasking, P., & St John, N. J. (2014). Prevalence of nonsuicidal self-injury in nonclinical samples: Systematic review, meta-analysis and meta-regression. Suicide and Life-Threatening Behavior, 44, 273–303. https://doi.org/10.1111/sltb.12070

Taliaferro, L. A., & Muehlenkamp, J. J. (2015). Risk factors associated with self-injurious behavior among a national sample of undergraduate college students. Journal of American College Health, 63(1), 40–48. https://doi.org/10.1080/07448481.2014.953166

Taylor, P. J., Jomar, K., Dhingra, K., Forrester, R., Shahmalak, U., & Dickson, J. M. (2018). A meta-analysis of the prevalence of different functions of non-suicidal self-injury. Journal of Affective Disorders, 227, 759–769. https://doi.org/10.1016/j.jad.2017.11.073

Tillman, K. S., Prazak, M., & Obert, M. L. (2018). Understanding the experiences of middle school girls who have received help for non-suicidal self-injury. Clinical Child Psychology and Psychiatry, 23(4), 514–527. https://doi.org/10.1177/1359104517743784

Valencia-Agudo, F., Burcher, G. C., Ezpeleta, L., & Kramer, T. (2018). Nonsuicidal self-injury in community adolescents: A systematic review of prospective predictors, mediators and moderators. Journal of Adolescence, 65, 25–38. https://doi.org/10.1016/j.adolescence.2018.02.012

Veldhuis, C. B., Nesoff, E. D., McKowen, A. L. W., Rice, D. R., Ghoneima, H., Wootton, A. R., Papautsky, E. L., Arigo, D., Goldberg, S., & Anderson, J. C. (2021). Addressing the critical need for long-term mental health data during the COVID-19 pandemic: Changes in mental health from April to September 2020. Preventive Medicine, 146, 106465. https://doi.org/10.1016/j.ypmed.2021.106465

Victor, S. E., Muehlenkamp, J. J., Hayes, N. A., Lengel, G. J., Styer, D. M., & Washburn, J. J. (2018). Characterizing gender differences in nonsuicidal self-injury: Evidence from a large clinical sample of adolescents and adults. Comprehensive Psychiatry, 82, 53–60. https://doi.org/10.1016/j.comppsych.2018.01.009

Vindegaard, N., & Benros, M. E. (2020). COVID-19 pandemic and mental health consequences: Systematic review of the current evidence. Brain, Behavior, and Immunity, 89, 531–542. https://doi.org/10.1016/j.bbi.2020.05.048

Waals, L., Baetens, I., Rober, P., Lewis, S., Van Parys, H., Goethals, E. R., & Whitlock, J. (2018). The NSSI Family Distress Cascade Theory. Child and Adolescent Psychiatry and Mental Health, 12, 52. https://doi.org/10.1186/s13034-018-0259-7

Wang, X., Hegde, S., Son, C., Keller, B., Smith, A., & Sasangohar, F. (2020). Investigating mental health of US college students during the COVID-19 pandemic: Cross-sectional survey study. Journal of Medical Internet Research, 22(9), e22817. https://doi.org/10.2196/22817

Whitlock, J., Muehlenkamp, J., Purington, A., Eckenrode, J., Barreira, P., Baral Abrams, G., Marchell, T., Kress, V., Girard, K., Chin, C., & Knox, K. (2011). Nonsuicidal self-injury in a college population: General trends and sex differences. Journal of American College Health, 59(8), 691–698. https://doi.org/10.1080/07448481.2010.529626

Wilkinson, P. O., Qiu, T., Neufeld, S., Jones, P. B., & Goodyer, I. M. (2018). Sporadic and recurrent non-suicidal self-injury before age 14 and incident onset of psychiatric disorders by 17 years: Prospective cohort study. British Journal of Psychiatry, 212(4), 222–226. https://doi.org/10.1192/bjp.2017.45

Wolff, J. C., Thompson, E., Thomas, S. A., Nesi, J., Bettis, A. H., Ransford, B., Scopelliti, K., Frazier, E. A., & Liu, R. T. (2019). Emotion dysregulation and non-suicidal self-injury: A systematic review and meta-analysis. European Psychiatry, 59, 25–36. https://doi.org/10.1016/j.eurpsy.2019.03.004

